# Fast ripples reflect increased excitability that primes epileptiform spikes

**DOI:** 10.1101/2023.03.26.23287702

**Authors:** Shennan A Weiss, Itzhak Fried, Jerome Engel, Michael R. Sperling, Robert K.S. Wong, Yuval Nir, Richard J Staba

## Abstract

The neuronal circuit disturbances that drive interictal and ictal epileptiform discharges remains elusive. Using a combination of extraoperative macro- and micro-electrode interictal recordings in six presurgical patients during non-rapid eye movement (REM) sleep we found that, exclusively in the seizure onset zone, fast ripples (FR; 200-600Hz), but not ripples (80-200 Hz), frequently occur <300 msec before an interictal intracranial EEG (iEEG) spike with a probability exceeding chance (bootstrapping, p<1e-5). Such FR events are associated with higher spectral power (p<1e-10) and correlated with more vigorous neuronal firing than solitary FR (generalized linear mixed-effects model, GLMM, p<1e-3) irrespective of FR power. During the iEEG spike that follows a FR, action potential firing is lower than during a iEEG spike alone (GLMM, p<1e-10), reflecting an inhibitory restraint of iEEG spike initiation. In contrast, ripples do not appear to prime epileptiform spikes. We next investigated the clinical significance of pre-spike FR in a separate cohort of 23 patients implanted with stereo EEG electrodes who underwent resections. In non-REM sleep recordings, sites containing a high proportion of FR preceding iEEG spikes correlate with brain areas where seizures begin more than solitary FR (p<1e-5). Despite this correlation, removal of these sites does not guarantee seizure freedom. These results are consistent with the hypothesis that FR preceding EEG spikes reflect an increase in local excitability that primes EEG spike discharges preferentially in the seizure onset zone and that epileptogenic brain regions are necessary, but not sufficient, for initiating interictal epileptiform discharges.

## Introduction

Spontaneous interictal epileptiform spikes (*i.e*., spikes) were first recognized nearly a century ago in extracellular recordings from animals exposed to chemoconvulsants and in the electroencephalogram (EEG) of patients with epilepsy^1^. Spikes are of primary importance in the diagnosis of epilepsy^2^, but also disrupt cognition^3^, and may, in some cases, promote the development of spontaneous seizures^4–6^. Epileptiform spikes, and their underlying neuronal correlates, are not uniform^5–7^. Moreover, spikes propagate as traveling waves^8,9^. Recent data suggests that epileptogenic brain regions may be identified by characterizing the spike initiation zone (SIZ) using precise source localization algorithms in intracranial EEG (iEEG)^9^. Little is known about what distinguishes spikes in the SIZ and what neuronal or macroscale neuronal network activity may prime an epileptiform discharge to occur next there.

High-frequency oscillations (HFOs) also are considered biomarkers of epileptogenic brain^10^. HFOs are subcategorized as fast ripples (FR, 200-600 Hz) and ripples (80-200 Hz). Like spikes FR, recorded by macroelectrodes, do not generally occur in healthy brain tissue^11^, but unlike spikes FR are thought to better predict the development of chronic seizures in chemoconvulsant exposed animals^12^, and better identify the epileptogenic zone (EZ), that is necessary and sufficient for seizure generation in patients undergoing evaluation for epilepsy surgery^13^. The physiological counterpart to FR is ripples that play a role in normal brain function, and specifically the sharp-wave ripple complex in area CA1 of the hippocampus has been found to be essential for memory consolidation during non-REM sleep^14^. However, in chemoconvulsant animal models and in patients with epilepsy, ripples are generated at increased rates in and around the EZ^15^. HFOs can occur superimposed on the background EEG (*i.e*., FR or ripple on oscillation [fRonO or RonO]) or superimposed on an interictal epileptiform spike (*i.e*., FR or ripple on spike [fRonS or RonS])^16,17^ and both are associated with ictogenesis in animal models^18,19^ and *in vitro*^6^ and *in vivo*^20^ recordings from epilepsy patients. However, fRonS and RonS may have increased accuracy for predicting epileptogenesis and localizing epileptogenic regions^17,21–24^. Recently, we reported that fRonO can also spatially propagate in epileptogenic regions within a range <30 mm at a velocity of ∼1.54 mm/ms^25,26^ and interestingly, propagation increased the probability a fRonO was immediately followed by a spike. These data suggest excitability associated with fRonO could help generate spikes, but the neuronal activity contributing to fRonO and ensuing spike has not been studied.

To better understand and possibly establish a link between fRonO events and after-going spike probability we investigated the neuronal and network activity associated with these events in patients with medically refractory focal epilepsy undergoing pre-surgical evaluation with paired macro- and microelectrode implants. We compared neuronal action potentials (AP) with the properties of HFOs and corresponding local iEEG spikes, as well as HFO-spike temporal coincidence (Figure 1). Our results show that in the SIZ, fRonO reflect increased neuronal excitability that precedes and promotes the generation of spikes restrained by inhibition. The putative SIZ almost always overlapped with epileptogenic regions, but not all epileptogenic regions were part of the SIZ as seizures often persisted after SIZ resection. These results bridge our understanding of clinically important interictal biomarkers of epilepsy, offer important new insights to the origins of pathological brain activity, and suggest that resecting the SIZ alone will not successfully control seizures.

**Figure 1:**
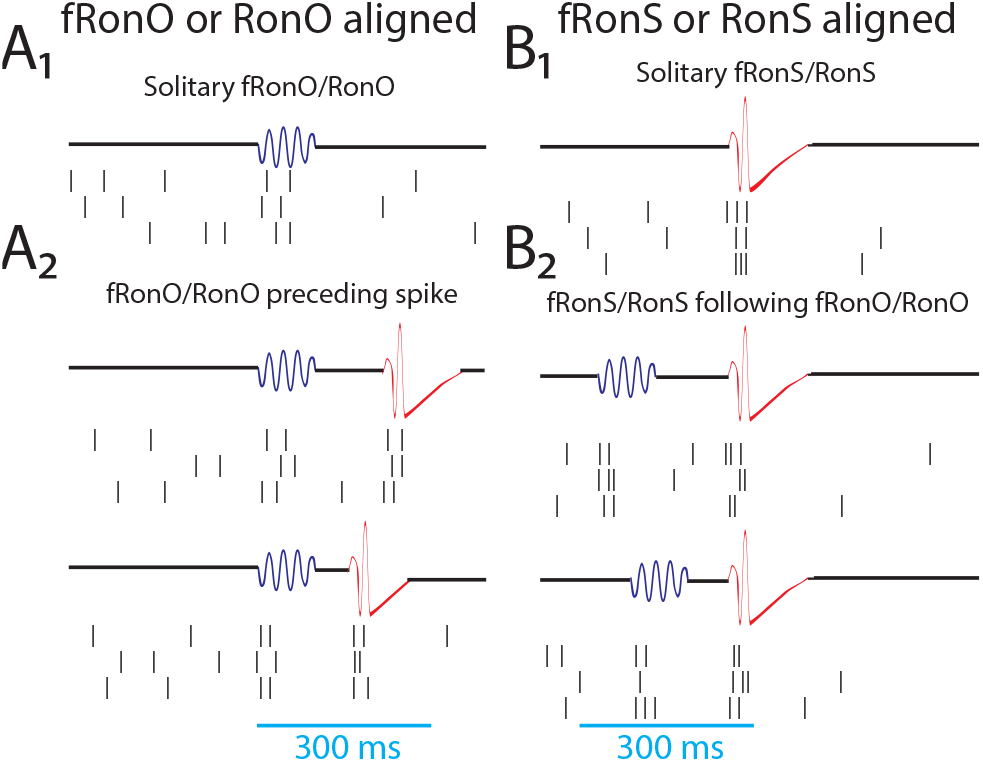
Schematic of the analytical methods comparing unit action potentials with fast ripples on oscillations (A,fRonO,blue), ripples on oscillations (A,RonO,blue), fast ripples on spikes (B,fRonS,red), and ripples on spikes (B,RonS,red), as well as the temporal coincidence of these HFO event types within 300 msec (A2,B2). Action potential trains were either temporally aligned to the onset for fRonO/RonO events (A) or the onset of fRonS/RonS events (B). Events were considered solitary if a fRonO or RonO did not precede a fRonS/RonS by <300 ms, and sharp-spikes, lacking an HFO, were also accounted for (A2). HFO event-unit trials aligned to fRonO/RonO events served to assess changes in excitability between solitary fRonO/RonO (A1) and the fRonO/RonO that preceded (<300 ms) spikes (A2). In contrast, HFO event-unit trials aligned to fRonS/RonS (B) assessed the changes in excitability between solitary fRonS/RonS (B1) and fRonS/RonS that followed (<300 ms) fRonO/RonO (B2).

## Methods

### Dataset Collection

All data were acquired with approval from the local institutional review board (IRB) at University of California Los Angeles (UCLA) and Thomas Jefferson University (TJU). The paired macroelectrode-microelectrode recording cohort was recorded from six patients with focal epilepsy at UCLA who were implanted for the purpose of localization of the SOZ in 2009-2010^27,28^. Sleep studies were conducted in the epilepsy monitoring unit 48-72 hr after surgery and lasted 7 hr on average, and sleep-wake stages were scored according to established guidelines. The montage included two EOG electrodes; two EMG electrodes scalp electrodes at C3, C4 Pz, and Fz; two earlobe electrodes used for reference; and continuous video monitoring. In each patient, 8–12 depth electrodes were implanted targeting medial brain areas. Both scalp and depth interictal intracranial EEG (iEEG) data, from the most medial depth electrode macroelectrode contact, were continuously recorded, during slow wave sleep, with a Stellate amplifier at a sampling rate of 2 kHz, bandpass-filtered between 0.1 and 500 Hz, and re-referenced offline to the mean signal recorded from the earlobes. Each electrode terminated in eight 40-μm platinum-iridium microwires from which extracellular signals were continuously recorded (referenced locally to a ninth noninsulated microwire) at a sampling rate of 28 kHz using a Neuralynx Cheetah amplifier and bandpass-filtered between 1 and 6000 Hz.

The retrospective resection cohort used consecutive recordings selected from 8 patients who underwent intracranial monitoring with depth electrodes between 2014 and 2018 at UCLA and from 15 patients at the Thomas Jefferson University (TJU) in 2016–2018 for the purpose of localization of the SOZ^29^. Inclusion criteria for this cohort included pre-surgical MRI for MRI-guided stereotactic electrode implantation, as well as a post-implant CT scan to localize the electrodes, and stereo EEG recordings during non-rapid eye movement (REM) sleep at a 2 kHz sampling rate, and a post-resection/ablation MRI. Patients with no adequate post-operative clinical follow up, or a failure to record at least ten minutes of artifact free iEEG during non-REM sleep were excluded.

Eligible patients were identified through queries of pre-existing clinical databases. Post-implantation CT scans were co-registered and normalized with the pre-implant and post-resection MRIs using Advanced Neuroimaging Tools (ANTs) (https://picsl.upenn.edu/software/ants/) with neuroradiologist supervision, using an in-house pipeline^30,31^. The position of each electrode contact was localized to normalized MNI coordinates. For each patient, in the resection cohort clinical interictal iEEG (0.1–600 Hz; 2000 samples per second) was recorded from 8 to 16 depth electrodes, each with 7–15 contacts, using a Nihon-Kohden 256-channel JE-120 long-term monitoring system (Nihon-Kohden America, Foothill Ranch, CA, USA) during epochs containing mostly high amplitude slow and delta oscillations. A larger number of electrodes with more contacts were implanted at TJU. The reference signal used for the recordings performed at UCLA was a scalp electrode position at Fz in the International 10–20 System. The reference signal used for the TJU recordings was an electrode in white matter.

### Spike sorting and characterization of high-frequency oscillations (HFOs) and epileptiform spikes

Action potentials were detected by high-pass filtering the local field potential (LFP) recordings above 300 Hz and applying a threshold at 5 SD above the median noise level. Detected events were further categorized as noise, single-unit, or multiunit events using superparamagnetic clustering^32^. Unit stability throughout sleep recordings was confirmed by verifying that spike waveforms and inter-spike-interval distributions were consistent and distinct throughout the night. HFOs and sharp-spikes were detected in the non-REM sleep iEEG using previously published methods^22,33,34^ (https://github.com/shenweiss). HFOs in the microwire LFP recording were not subject to analysis, minimizing the chance that high-frequency events such as fast ripples may be influenced by leakage from action potential events^35^. A two-stage algorithm first used a custom Hilbert based detector implemented in Matlab with artifact rejection features to identify transient elevations in ripple (80-200 Hz) and fast ripple (200-600 Hz) amplitude^33^. In the second stage, the different HFO types: (1) ripples on oscillations (RonO); (2) ripples on spikes (RonS); (3) fast ripples on oscillations (fRonO); (4) fast ripples on spikes (fRonS), and (5) sharp-spikes without a HFO were distinguished using the topographical analysis of the wavelet convolution^22,34,36^. In brief, this method distinguished HFOs on spikes by comparing onset of the outermost closed-loop isopower contour of the HFO in the time-frequency spectrogram with onset of the outermost open-loop isopower contour of the spike and determining if the latter occurred earlier. Open-loop contours without closed loop contours were designated sharp-spikes without HFOs. This method also identified the spectral content, power, and duration of each HFO. Ripples and fast ripples were distinguished by the mean frequency of the closed-loop contour group, and peak spectral content and power from the largest isopower contour value in the closed-loop contour group. Duration was computed from the onset and set of the outermost contour in the closed-loop contour group. Compared to visual analysis of two reviewers, HFO and spike detection sensitivity and accuracy of event detection, using this method, was typically 80–90%^33,34^. Following automatic detection of HFO and sharp-spikes, false detections of clear muscle and mechanical artifact were deleted by visual review in Micromed Brainquick. This methodology did not detect bluntly contoured spikes, with no corresponding power signature >80 Hz, and sharp-waves.

To calculate the latency, within individual iEEG channels, between HFOs on oscillations and after-going epileptiform spikes, the onset times of all spikes, or a subtype of spikes, were subtracted from the onset time of each individual HFO on oscillation to find the minimum latency greater than zero. To determine if the distribution of these latencies exceeded chance, the number of HFO on oscillations were kept constant but the onset times of the HFOs on oscillations across the recording time were uniformly randomized within artifact free epochs and resampled 500 times within each individual iEEG channel and across all iEEG channels.

### Preparing and implementing generalized linear mixed-effects models (GLMMs)

All the RonO, fRonO, RonS, or fRonS events detected in a single macroelectrode contact’s recording were individually compared with the corresponding action potentials (APs) from each individual unit isolated from the LFPs recorded from the bundle of microelectrodes distal to that macroelectrode (within <∼4mm). Within each individual unit, for each macroelectrode HFO event, a trial was generated consisting of a two second raster centered at the onset of the HFO event with a resolution of 1 ms (Figure 1,2A). This raster was then convolved with a 100 ms Gaussian kernel and the resulting AP train was down sampled to 40 Hz. We used a 100 ms kernel to better quantify mean HFO related firing as opposed to examining fine temporal structure. For each HFO event-unit trial, the pre-event baseline unit firing rate was defined as the mean firing rate of the Gaussian smoothed AP train rate beginning 750 msec prior to HFO onset until the event (*i.e*., bl-fr). The mean was used, as opposed to the median, to avoid bias towards the isolated single units with low firing rates. The peak HFO-unit firing rate was defined as the maximum of the Gaussian smoothed AP train rate during the duration of the HFO event (*i.e*., hfo-fr), and another value hfodiff-fr was defined as hfo-fr minus bl-fr. During RonS and fRonS the peak firing rate during the epileptiform spike was invariably during the superimposed HFO^17^. Since we wished to examine the factors that reliably modulate AP firing during HFO events, we excluded units from the model that did not significantly change in firing rates during the HFO events. Often this occurred because few HFO events were detected in the unit’s corresponding macroelectrode recording. Thus, a paired t-test was used to compare the bl-fr with the hfo-fr for each type of HFO event, across all trials, but within units. If the p value exceeded 0.001 after controlling for the Holms-Bonferroni false-discovery rate (f.d.r.) the unit was excluded from the generalized linear mixed-effects models (GLMMs) analysis.

**Figure 2:**
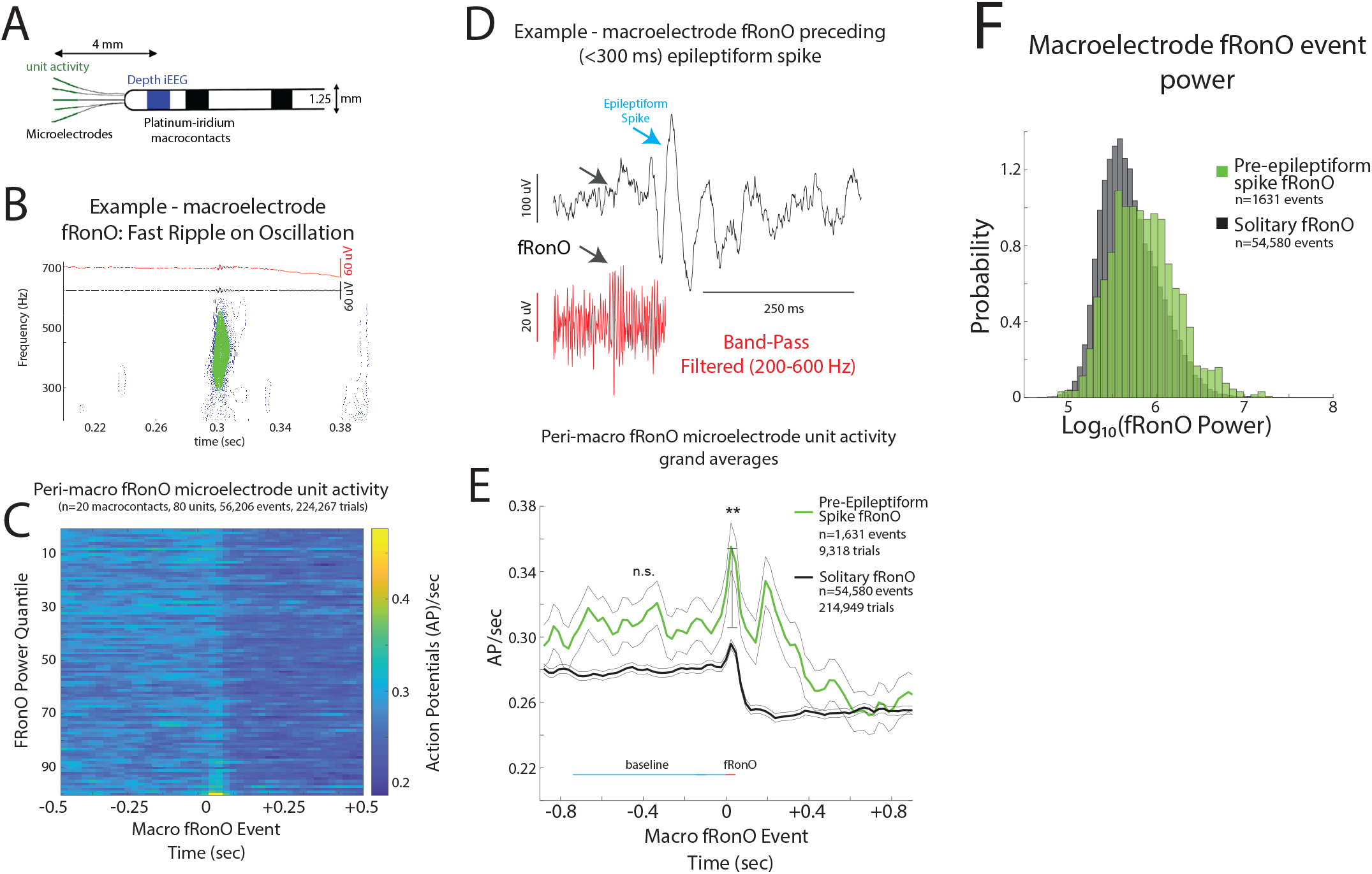
iEEG fast ripple on oscillation (fRonO) preceding (<300 ms) an interictal iEEG spike are associated with higher spectral power and higher unit firing rate. (A) An illustration of a Behnke-Fried depth electrode. The HFO and spike events are detected in the iEEG recorded from the mesial most depth macroelectrode contact shown in blue, while the unit activity associated with these HFO and spike events is isolated from the local field potentials recorded from the microelectrodes in green. Comparisons between the HFO and spike events and unit activity were made exclusively within a single Behnke-Fried depth electrode, and not across different depth electrodes. (B) An example fRonO identified using the topographical analysis of the wavelet convolution that quantifies event power. (C) Peri-fRonO unit gaussian smoothed action potential (AP) firing rates from 80 units, that exhibited a significant fRonO triggered increase in firing (p<0.001, f.d.r. corrected), averaged by quantiles defined by the log10(power) of the fRonO events. Higher power fRonOs are associated with a greater increase in AP firing rate (GLMM, p<<e-10, Table S2), and AP firing rate decreases after the fRonO event. (D) Representative example of a fRonO event preceding a spontaneous epileptiform discharge with after-going slow wave in a macroelectrode recording. (E) Grand average of the gaussian smoothed fRonO event-unit AP train trials for solitary peri-fRonO event trials not preceding an epileptiform discharge (black, 95% confidence interval), and peri-fRonO event trials that did precede (<300ms) an epileptiform discharge (green). fRonO events that preceded an epileptiform discharge, relative to those that did not, had higher peak AP firing rates during the fRonO (red line) compared to baseline (cyan line) (**, GLMM, p<1e-3). (F) Normalized histogram of fRonO event spectral power. fRonO events preceding spikes had a larger power (t-test, p<1e-10, Cohen’s d=.475) than solitary fRonO, but the interaction of fRonO power with pre-spike status did not explain the increase in firing rates (GLMM) during fRonO preceding spikes.

The data for the GLMMs consisted of all the combined HFO event-unit AP train trials across all units, macroelectrodes, and patients. For each category of HFO, the GLMMs fit the bl-fr or the hfodiff-fr using a Gaussian distribution, a log link, a fixed intercept, and the maximum quasi-likelihood method in Matlab using the fitglme.m function. Random effects included the unit identifier, macroelectrode contact identifier, and patient identifier. All these identifiers were distinct (*i.e*., unit numbers did not repeat across different macro-microelectrode pairs). Fixed effect included the log10(spectral power of the HFO event) recorded in the macroelectrode, and whether the RonO or fRonO preceded (<300 ms) an epileptiform spike or whether a RonS or fRonS followed (<300 ms) a RonO or fRonO in the macroelectrode recording. At most, each GLMM included only two fixed effects, including HFO power, with a single interaction term. The categorical neuroanatomical location of the unit was not used as a fixed effect because the unit identifier was included as a random effect.

### Identification of the seizure onset zone (SOZ) and resected electrodes and related metrics

With respect to seizure outcome following surgery, Engel class 1 refers to freedom from disabling seizures, Engel class 2 to rare disabling seizures, Engel class 3 to worthwhile improvement in seizures, and Engel class 4 to no worthwhile improvement. The epileptologist defined SOZ contacts were aggregated across all seizures during the entire iEEG evaluation for each patient. The identification of the named electrode contacts in the resection cavity was performed manually in itk-SNAP (https://itk-snap.org) prior to performing calculations^29^. The SOZ and resected ratio was calculated, within patients, as the number of HFO events detected from the SOZ or resected contacts, respectively, divided by the total number of detected HFO events. The fast ripple rate-distance radius difference was calculated by first deriving the rate (events/min) of all fRonS events and the fRonO events with a peak spectral frequency > 350 Hz per contact and then computing an adjacency matrix as the average rate between contact pairs multiplied by the Euclidian distance (mm) between the contact pairs. A second adjacency matrix was computed using only contacts in the resection margins. The charpath.m function (BCT, https://sites.google.com/site/bctnet/) was used to calculate the radius of these scaled distance graphs and the radius difference was defined as the √(radius of all electrodes-radius of the resected electrodes)^29^.

## Results

### Patient characteristics and isolation of electrophysiological events

To study the intricate relations between HFOs, interictal spikes and neuronal spiking activity, we combined microelectrode and macroelectrode data recorded during NREM sleep, as well as post-surgical seizure outcome data in two separate patient cohorts. First, in six patients with medically refractory focal seizures who were implanted with depth electrodes, 3 were diagnosed with mesial-temporal lobe epilepsy (MTLE), one with MTLE and neocortical epilepsy, one with neocortical epilepsy, and one with an unknown site of seizure onset. Each of the depth electrodes in these patients contained a bundle of 8 microelectrodes extending beyond the distal tip that was positioned in mesial temporal lobe or medial neocortical structures (Figure 2A)^27^. We isolated HFOs and sharp-spikes in the iEEG and action potentials (APs) from 217 units in 175.95 iEEG contact-hours of synchronized macroelectrode and microelectrode recordings during N2/N3 sleep (Table 1). Second, to verify the findings in this small cohort had clinical significance, we isolated HFOs and sharp-spikes in 1009.7 contact-hours of iEEG recordings during non-REM sleep in a separate cohort of 23 patients who underwent the same invasive EEG tests, but were not implanted with microelectrodes, and who subsequently had resections for their medically refractory focal seizures (Table S1)^29^.

**Table 1:** Clinical demographics and a full description of the brain structures and number of units identified in the recordings. Abbreviations: L, left hemisphere; R, right hemisphere; H, hippocampus; A, amygdala; EC, entorhinal cortex; PHG, parahippocampal gyrus; TG, temporal gyrus; AC, anterior cingulate; MC, middle cingulate; PC, posterior cingulate; OF, orbitofrontal and medial prefrontal cortex; SM, supplementary motor area; P, parietal cortex; TO, temporo-occipital, PT, posterior temporal cortex; FG, fusiform gyrus; IFG, inferior frontal gyrus.

| Patient | PET/MRI Findings | Seizure onset zone (SOZ) | Resection/ Outcome | Macroelectrode/units |
| --- | --- | --- | --- | --- |
| 1 | Metabolic and structural abnormalities left temporal. Pathology FCD IB | Left temporal spreading to right entorhinal cortex | Left temporal, significant improvement | L H : , L A : , L PHG : 6 units, L AC : 9 units, L TO : , R H : 5 units, R A : , R EC : 11 units, R PHG : 2 units, R AC : , R TO : |
| 2 | Mild metabolic abnormality left temporal, structural abnormality right frontal lobe and left hippocampal T2 hyperintensity | Left medial temporal lobe | No surgery | L H : 6 units, L A : 8 units, L PHG : 13 units, L AC : 3 units, L SMA, R H : 8 units, R A : 8 units, R PH : 6 units, R AC : 4 units, R OF, R SMA |
| 3 | Mild metabolic abnormality right temporal, mild structural abnormality left temporal | Bilateral mesial temporal lobe and right inferior frontal gyrus | No surgery | L H : 3 units, L A : , L EC : 1 unit, L AC : 3 units, L OF : 10 units, R H : 16 units, R A : 8 units, R EC : 7 units, R TG : 12 units, R AC : , R OF : , R IFG : |
| 4 | Metabolic abnormality right parietal lobe, normal structural. Pathology FCD IIA & IIB | Right parietal | Right parietal seizure free | L H : , L A : , L PHG : 4 units, L P : 8 units, R H : 9 units, R A : , R PH : 8 units, R P : 7 units |
| 5 | Volume loss due to prior LAMTL. Normal around margins of resection site and elsewhere | Not localized | No surgery | L PH : 8 units, L AC : 1 unit, L OF : 11 units, L SMA : 8 units, L PT : 2 units, R H : 5 units, R PH : 6 units, R OF : 7 units, R P : 10 units |
| 6 | Metabolic and structural abnormalities left temporal including amygdala T2 hyperintensity. Pathology gliosis | Left mesial temporal lobe | Left temporal seizure free | L H : 5 units, L A : , L EC : 8 units, L OF : 7 units, R H : 1 units, R A : , R EC : 11 units, R OF : |

Many of these patients had predictors for poor post-surgical seizure outcome, including six who had normal MRI, 5 had multilobar SOZs, and 6 who were reoperated for a previous poor seizure outcome. Consequently, only 10 of these 23 patients were rendered seizure free (Table S1).

### Fast ripple on oscillation (fRonO) and associated neuronal firing are increased when followed by interictal spike

Based on results from our prior investigation of fRonO and epileptiform spikes we defined fRonO and RonO preceding spikes as occurring within <300 ms^26^. We defined iEEG spikes as either fRonS, RonS, or sharp-spikes (no fast ripple or ripple) which accounted for 19.8, 71.5, and 8.7% of all spikes, respectively. We compared peak firing during the fRonO and RonO events to a baseline epoch defined as beginning 750 ms before the events. We then asked if there was differential AP firing accompanying fRonO and RonO that preceded spikes versus solitary fRonO and RonO, i.e., without subsequent spike. Figure 1A provides a schematic illustration of the different categories used for this data analysis.

Of the 217 units, 80 and 174 increased their firing rate during the fRonO and RonO (t-test, p<1e-3, f.d.r. corrected), respectively, irrespective of coincidence with spikes, and no units decreased their firing rate during either HFO. Among the aforementioned units that increased firing during HFO with respect to baseline, we found higher peak firing rates, with respect to baseline, correlated with higher HFO power (log10 of spectral power) of all fRonO (GLMM, p<<1e-10, Table S2, Figure 2C) and all RonO power (GLMM, p<<1e-10, Table S4, Figure 3B). By contrast, lower baseline unit (baseline) firing rate 750 ms before the HFO correlated with higher power of all fRonO (GLMM, p<1e-6, Table S3) and all RonO power (GLMM, p<1e-10 Table S5).

**Figure 3:**
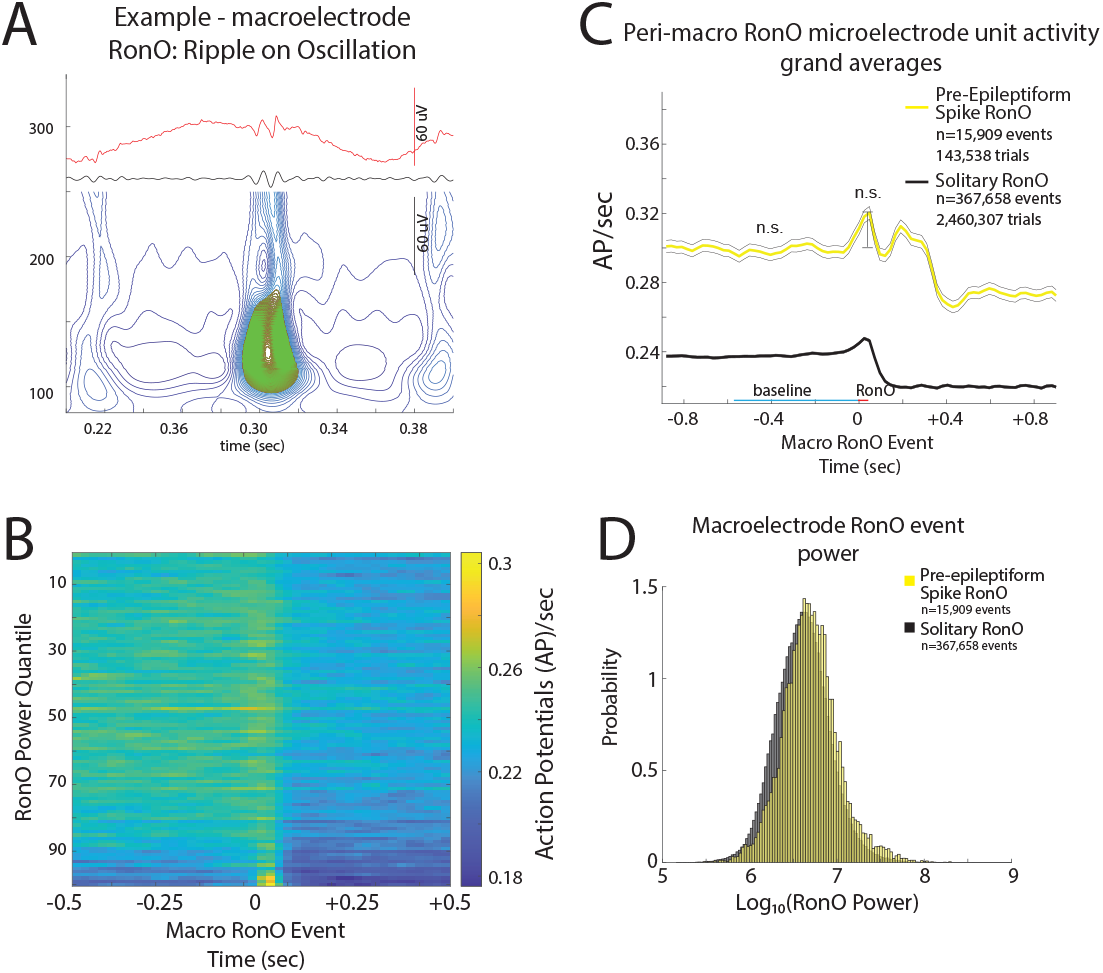
iEEG ripple on oscillations (RonO) associated increases in unit firing rate are not larger if the RonO precedes (<300 ms) an epileptiform spike. (A) An example RonO identified using the topographical analysis of the wavelet convolution. (B) Peri-RonO unit gaussian smoothed action potential (AP) firing rates from 174 units, that exhibited a significant RonO triggered increase in firing (p<0.001, f.d.r. corrected), averaged by quantiles defined by the log10(power) of the RonO events. Higher power RonOs are associated with a greater increase in AP firing rate (GLMM, p<e-10, Table S4), and AP firing rate decreases after the RonO event. (C) Grand average of the gaussian smoothed HFO event-unit AP train trials for all peri-RonO event trials not preceding an epileptiform discharge (black, dotted lines=95% confidence interval), and peri-RonO event trials that did precede (<300ms) an epileptiform discharge (yellow). Although the baseline (cyan line) firing rate appears different between the two conditions, no increase was detected in the GLMM that accounted for the random effects of patient, macroelectrode, and unit (GLMM, Table S5). Also, RonO events that preceded an epileptiform discharge, relative to those that did not, had a decreased peak AP firing rate during the RonO event (red line) compared to baseline (cyan line)(GLMM, p<5e-3, Table S4). (D) Normalized histogram of fRonO event power in macroelectrode recordings. RonO events preceding spikes had a larger power (t-test, p<1e-10, Cohen’s d=0.3) than solitary RonO and the interaction of RonO event power and pre-spike status significantly predicted increased firing rate (GLMM, 1e-4, Table S4).

We next examined whether neuronal firing was modulated depending on whether the fRonO/RonO preceded (<300 ms) a iEEG spikes (Fig 1A). Baseline firing rate was not statistically different for fRonO preceding spikes compared to firing during the solitary fRonO (GLMM, n.s., Table S3, Figure 2E). A marginally significant reduction in baseline firing rate was present for RonO preceding spikes (GLMM, p<0.05, Table S5). Following the solitary fRonO or RonO, unit firing rate decreased relative to baseline (Figure 2C,2E,3B,3C). However, when a fRonO or RonO preceded a spike (*i.e*., positive pre-spike status, Figure 2D) the gaussian smoothed HFO event-unit AP train trials exhibited a second peak in firing rate following the HFO event (Figure 2E, 3C) corresponding to the spike. fRonO preceding spikes had greater spectral power compared to solitary fRonO (t-test, p<1e-10, Cohen’s d=.475, Figure 2F). During the fRonO, the peak firing rate relative to baseline was greater for fRonO preceding spikes than the firing during a solitary fRonO. This effect was driven by the “pre-spike” status of the fRonO (GLMM, p<5e-3, Table S2, Figure 2E,F), and not the interaction of increased power with pre-spike status which was found to decrease the spike rate (GLMM, p<1e-4, Table S2). Since fRonO power was increased for the pre-spike fRonO events, we analyzed pre-spike fRonO power at latencies between 0 and 300 ms preceding the spike. We found greater power throughout this range of latencies, but especially for fRonO that preceded spikes by <10 ms (Fig. S1).

When a RonO preceded a spike, the RonO spectral power was higher than the power for a solitary RonO, but the effect size was smaller than fRonO preceding a spike (t-test, p<1e-10, Cohen’s d=0.3, Figure 3D). Peak firing rate relative to baseline for RonO preceding spikes was decreased compared to the firing during solitary RonO based on pre-spike status alone (GLMM, p<5e-3, Table S5, Figure 3C), however the interaction of RonO power with pre-spike status significantly increased firing rate (GLMM, p<1e-4, Table S4). These results show that a fRonO, but not a RonO, preceding a spike reflects increased excitability.

### Inter-ictal fast ripple on spike (fRonS) and ripple on spike (RonS) associated neuronal firing are decreased when preceded by a fast ripple on oscillation (fRonO)

In this analysis, we shifted from fRonO/RonO aligned trials of AP trains to HFO spike (*i.e*., fRonS/RonS) aligned trials (Fig 1B). We analyzed peak AP firing during fRonS (Fig 4A) or RonS (Fig 4D), as compared to the 750 ms pre-event baseline epoch, and asked whether the extent of AP firing was different when fRonS/RonS was preceded (<300 ms) by fRonO or RonO than when fRonS/RonS occurred alone (Fig 1B). Of the 217 units, irrespective of coincidence with fRonO or RonO, 49 and 109 increased their firing rate during a fRonS and RonS, respectively (t-test, p<0.001, f.d.r. corrected). In contrast to a prior study of HFO spikes^17^, no units decreased their firing rate.

**Figure 4:**
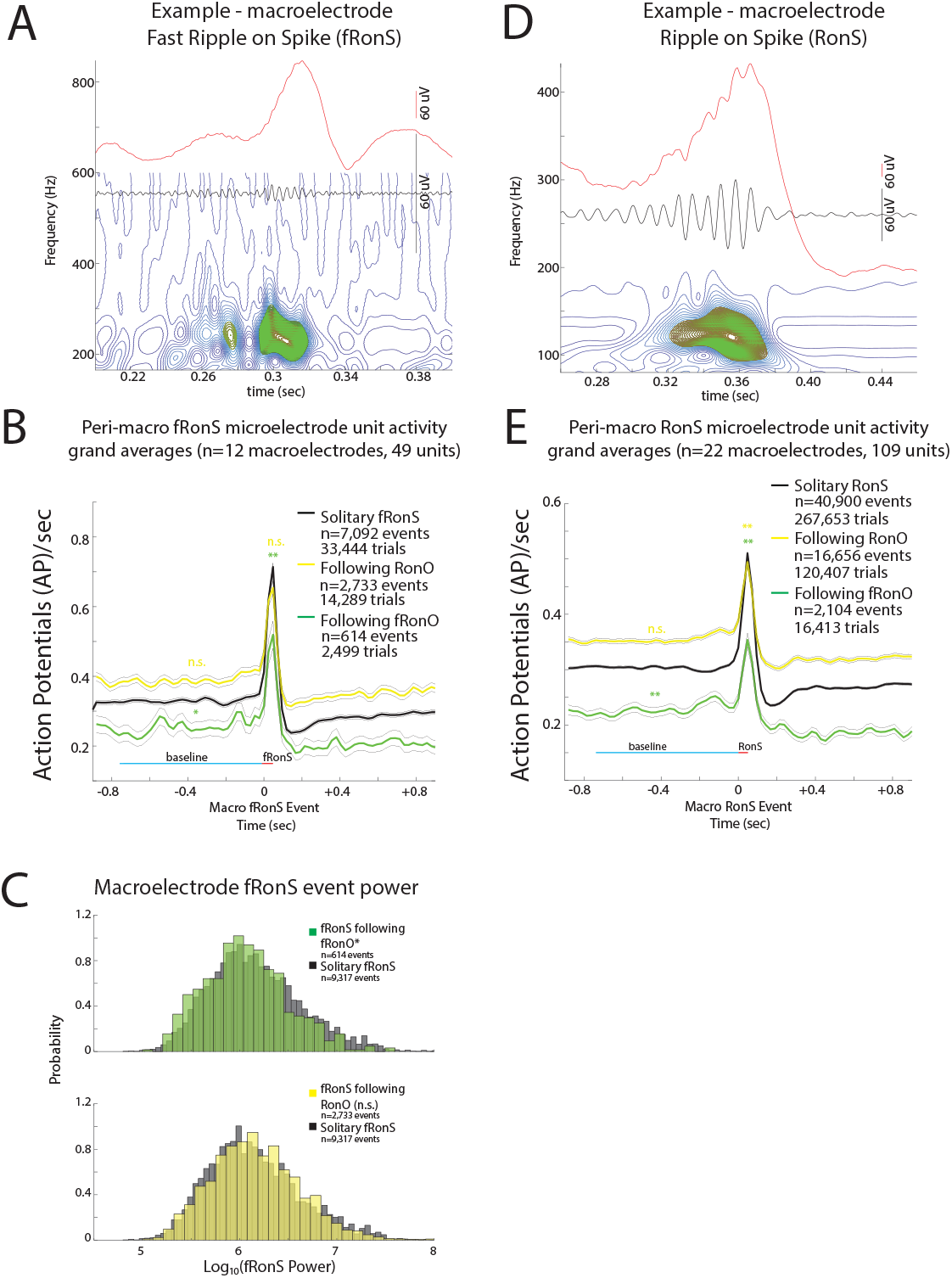
Unit firing rates rates prior to and during fast ripple on spike (fRonS) and ripple on spike (RonS) are decreased if preceded (<300 ms) by a fast ripple on oscillation (fRonO). An example fRonS (A) identified using the topographical analysis of the wavelet convolution. (B) Grand average of the gaussian smoothed HFO event-unit action potential (AP) train trials peri-fRonS from 49 units, that showed statistically significant increased fRonS triggered firing (p<0.001, f.d.r. corrected). Shown are trial averages with no preceding event (black, neither ripple on oscillation [RonO] or fRonO), when a fRonS followed a ripple on oscillation (RonO, yellow), or when a fRonS followed a fRonO (green). AP firing rate during the fRonS was proportional to fRonS log10(power) (GLMM, p<1e-10, not shown, Table S6). The baseline AP firing rate (cyan line) for fRonS that followed a fRonO was significantly lower than fRonS event without a preceding event (*, GLMM, p<0.05, Table S8), while fRonS that followed a RonO had no change in baseline AP firing rate (n.s., GLMM, Table S9). During the fRonS event (red line) the peak AP firing rates relative to baseline (cyan line) was decreased when fRonS events followed a fRonO (**, GLMM, p<1e-10, Table S6), but no decrease, compared to solitary fRonS, was seen if a fRonS followed a RonO. (C) Normalized histogram of fRonS event power in macroelectrode recordings. fRonS power was reduced when it followed a fRonO (t-test, p<1e-5, Cohen’s d=0.21) but unchanged if the fRonS followed a RonO. The interaction of fRonS event power and pre-fRonO status, but not pre-RonO status, was significant (GLMM, p<1e-10) in predicting firing rate (Table S6,S7). (D) An example RonS identified using the topographical analysis of the wavelet convolution. (E) Grand average of the gaussian smoothed HFO event-unit AP train trials rates peri-RonS from 109 units, that showed statistically significant increased RonS triggered firing (p<0.001, f.d.r. corrected) as in (B). The baseline AP firing rate (cyan line) for RonS that followed a fRonO was significantly lower than RonS event without a preceding event (**,GLMM, p<1e-13), while RonS that followed a RonO had no change in baseline AP firing rate (Table S12,S13). During the RonS event (red line) the peak AP firing rates, relative to baseline (cyan line), was decreased when RonS events followed either a fRonO or a RonO (**, p<1e-10).

Among all fRonS and RonS, irrespective of preceding fRonO/RonO, we found higher peak firing rates, with respect to baseline, correlated with higher HFO power (log10 of spectral power) of fRonS (GLMM, p<1e-10, Table S6,S7) and RonS power (GLMM, p<<1e-10, Table S10,11). Also, during the baseline epoch beginning 750 ms before the fRonS/RonS, slower firing rate correlated with higher spectral power of RonS (GLMM, p<1e-10, Table S12,S13), and fRonS (GLMM, p<0.05, Table S9).

When we compared the fRonS that followed fRonO with the solitary fRonS it confirmed that the baseline firing rate, fRonS power, and the corresponding peak unit firing rates, were all reduced in the former. Specifically, baseline firing rate was lower if a fRonS followed a fRonO (*i.e*., positive followingFRonO status, GLMM, p<0.05, Table S8, Figure 4B) but baseline firing rate was unchanged if a fRonS followed a RonO (*i.e*., positive following RonO status, GLMM, p>0.05, Table S9, Figure 4B). Spectral power was lower for fRonS following fRonO (t-test, p<1e-5, Cohen’s d=0.21, Figure 4C), but unchanged for fRonS following RonO (t-test, p>0.05, Figure 4C). Compared with solitary fRonS, peak firing rate during the fRonS, relative to the 750 ms pre-event baseline, was decreased for fRonS that followed a fRonO, but not if the fRonS followed a RonO (GLMM, p>0.05, Table S7, Figure 4B), and this effect was driven by the “followingfRonO” status (GLMM, p<1e-5, Table S6, Figure 4B), and the interaction of lower fRonS power and “followingfRonO” status (GLMM, p<1e-5, Table S6, Figure 4C). To better understand this effect, we examined fRonS that followed fRonO power between 0 and 300 ms and found that fRonS power was only decreased for events that occurred at latencies >10 ms (Fig S2). This indicates that fRonS that follow fRonO by <10 ms may be distinct from fRonS that follow fRonO by 10-300 ms (Fig S2).

When we compared the RonS that followed fRonO with the solitary RonS it also confirmed that the baseline firing rate and the corresponding peak unit firing rates, were reduced in the former. In detail, baseline firing rate was reduced when a RonS followed a fRonO (GLMM, p<1e-10, Table S12, Figure 4E) but not when a RonS followed a RonO (GLMM, p>0.05, Table S13, Figure 4E). RonS spectral power was unchanged for RonS following fRonO but was smaller for RonS following RonO (t-test, p<1e-10, Cohen’s d=0.22). Peak firing rate during the RonS, relative to baseline, was decreased when the RonS followed a fRonO which was driven exclusively by “followingfRonO” status (GLMM, p<1e-10, Table S10, Figure 4E). However, in the case of RonS following RonO a decrease in peak firing was also observed that was driven by both “followingRonO” status (GLMM, p<1e-6, Table S11, Figure 4E) and the interaction of “followingRonO” status with lower RonS power (GLMM, p<1e-6, Table S11).

In summary, Figure 4 shows when a fRonS/RonS follow a fRonO the excitability during and before the fRonS/RonS is decreased.

### fRonO preceding interictal spikes demarcate a spike initiation zone in epileptogenic tissue

Thus far we found increased excitability during fRonO and decreased excitability during the ensuing fRonS and RonS. Next we quantified where fRonO and fRonS/RonS occur and whether the location might correspond with a spike initiation zone (SIZ). We assumed that if fRonO prime fRonS or RonS, then rate of fRonO should exceed the rate of fRonS or RonS in the SIZ. To test this we calculated the ratio of fRonS to fRonO and RonS to fRonO occurrences in one-minute bins in the SOZ. We found the ratio fRonS to fRonO was less than one, but RonS to fRonO was greater than one (Figure 5A). We next examined whether fRonO preceding fRonS or RonS occurred more often than chance using boot strapping (see Methods). In the SOZ, the probability of a fRonO preceding fRonS or RonS (<300 ms) was greater than chance (n=500 surrogates, z score=4.61, p<1e-5, Figure 5B), but in the non-SOZ, fRonO preceding fRonS or RonS was less than chance (z=-7.72, p>0.05, Figure 5B). Specifically, in the SOZ, greater differences between the observed values and boot strapped-surrogates were found with fRonO preceding fRonS (z=11.61, p<<1e-10, Figure 5B) than the differences for fRonO preceding RonS (z=3.63, p<1e-4, Figure 6B). In contrast to fRonO, in the SOZ and non-SOZ, RonO preceding fRonS or RonS occurred less than chance (z=-15.44, p>0.05 and z=-7.75, p>0.05 respectively). Collectively these results indicate fRonO and not RonO likely prime spikes, especially fRonS in a SIZ.

**Figure 5:**
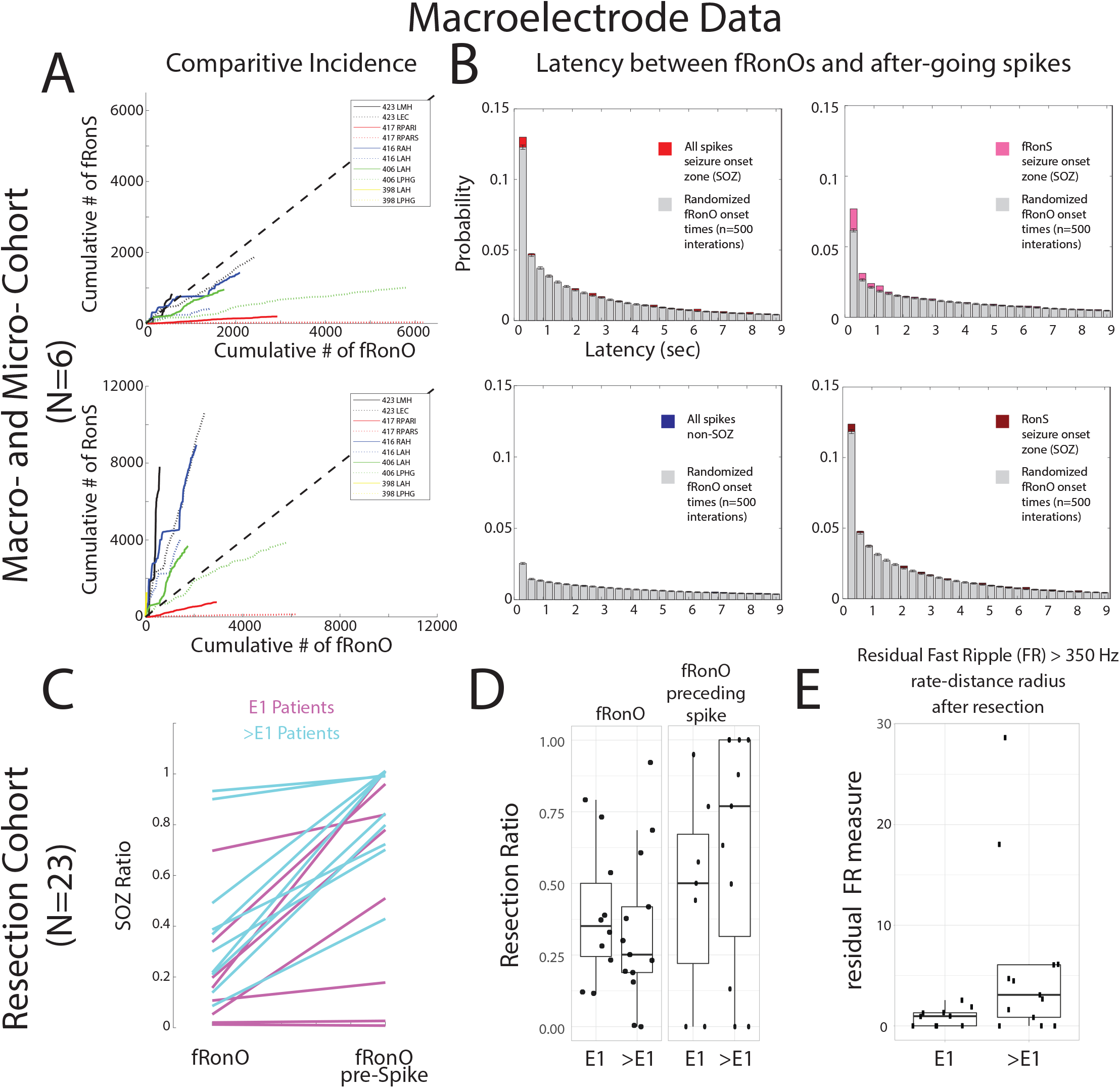
Fast Ripples on Oscillations (fRonO) preceding (<300 msec) epileptiform spikes occur mostly in the seizure-onset zone (SOZ), however resection of cortical territory generating fRonO with after-going spikes does not correlate with seizure freedom. In the paired macro- and microelectrode cohort, (A) Comparative incidence of fRonO with fast ripples on spikes (fRonS, top), as well as fRonO with ripples on spikes (RonS, bottom) in 10 macroelectrode channels in the SOZ calculated in bins of 60 seconds over the recording duration. Note that for most channels multiple fRonO occur before a fRonS is detected. However, RonS are generated in greater number than fRonO. (B) Normalized histogram of the latency between spike onset and fRonO in individual iEEG channels when computed using all spikes in the SOZ (red bars, n=41,861 fRonO), all spikes in the non-SOZ (blue bars, n=52,136 fRonO), only fRonS in the SOZ (pink bars), and only RonS in the SOZ (dark red bars). Latencies computed with boot-strapping statistics (see methods, n=500 surrogates, grey bars) indicated that fRonO in the <300 msec bin exceeded chance (z score=4.61, p<1e-5), but were less than chance in the non-SOZ (z=-7.72, p>0.05, blue). The most significant effect was seen for fRonO with after going fRonS (z=11.61, p<<1e-10). (C-E) Data taken from a cohort of 23 patients with stereo-EEG recordings, but no microelectrodes placed, who underwent resections. (C) The within-patient differences in the proportion of events in the SOZ for fRonO events compared to fRonO events preceding (<300 ms) spikes for patients with a seizure-free Engel 1 (E1) outcome (magenta), and non-seizure free patients (cyan, >E1, Engel 2-4). The proportion of fRonO events preceding spikes in the SOZ was significantly higher (ranksum, p<1e-5) (D) Box plots of the ratio of fRonO (left) and fRonO preceding spike (right) events within the resection margins stratified by post-operative seizure outcome. (E) A spatial graph theoretical measure, the fast ripple rate-distance radius resection difference, that quantifies both the volume and activity of the residual fast ripple generating tissue remaining after resection that was significantly decreased in seizure free patients (ranksum, p<0.05). Abbreviations L: left hemisphere R: right hemisphere AH/MH: hippocampus, EC: entorhinal cortex, PAR: parietal (sup./inf.), PHG: parahippocampal gyrus

**Figure 6:**
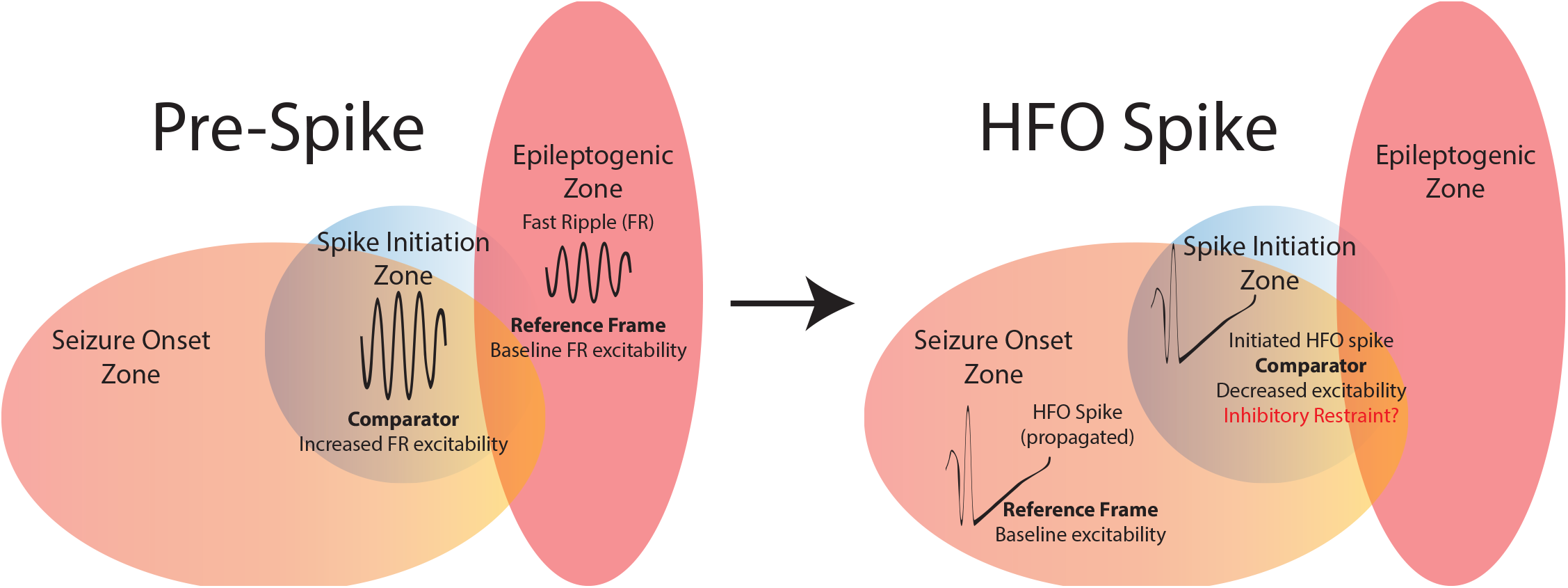
Schematic of the summary of findings. Resecting fast ripples on oscillations (fRonO) preceding spikes did not result in seizure freedom, thus the spike initiation zone (SIZ, blue) does not always strongly overlap with the epileptogenic zone (EZ, red). However, fRonO preceding spikes were found in greater proportions in the seizure onset zone (SOZ), so the SIZ does strongly overlap with the SOZ. Prior to spike initiation (right) a fRonO in the SIZ is generated with increased single unit firing rate relative to a smaller power FR in the EZ. Within 300 milliseconds of the FR occurring in the SIZ a fRonS or RonS in the SIZ is generated, suggesting the fRonO may facilitate the fRonS or RonS (HFO spike, right). Unit firing rate during an HFO spike in the SOZ, outside the SIZ, is relatively greater compared to the unit firing rate during the fRonS in the SIZ. The decreased excitability during the fRonS in the SIZ may be due to an inhibitory restraint mechanism that ordinarily prevents HFO spike initiation.

Extending the results from the preceding paragraph we assessed fRonO preceding all spikes (fRonS, RonS, sharp-spikes) in relation to the clinical SOZ and seizure outcome in a separate cohort of 23 patients who underwent resections. For each patient we computed the SOZ ratio defined as the proportion of fRonO preceding spikes on macroelectrode contacts in the SOZ contacts to the total macroelectrode contacts in SOZ and non-SOZ.

The SOZ ratio of fRonO preceding spikes was greater than the SOZ ratio of solitary fRonO (ranksum, p<1e-5, Figure 5C), but fRonO preceding spikes were not identified in five patients. In addition, across all patients, the proportion of fRonO preceding spikes in the SOZ was 66.5% and only slightly greater than solitary spikes at 64.5%. These results suggest fRonO preceding spikes and solitary HFO-spikes and sharp-spikes, but not solitary fRonO, occur mostly in the SOZ^37^.

Theoretically, resection of fRonO preceding spikes should correlate with good seizure outcome because it is found in high proportions in the SOZ. We found that the mean proportion of fRonO events recorded from contacts within the resection margins relative to the total number of fRonO events recorded from all the contacts [resection ratio] was higher across seizure free patients compared to those who were not seizure free. However, when we calculated these same resection ratios using fRonO preceding spikes the mean resection ratios were paradoxically lower in patients seizure free than those who were not seizure free, though differences were not statistically significant (ranksum, p>0.05, Figure 5D). Also, in all patients, we found a higher resection ratio of fRonO preceding spikes than solitary fRonO (signrank, p<0.05, Figure 5D), indicating that fRonO preceding spikes to a greater extent than solitary fRonO demarcate the clinical SOZ that was targeted for resection.

Since neither of the two fRonO resection ratios was significantly greater in the seizure free patients it raises the issue of whether fast ripples are satisfactory biomarkers of epileptogenic tissue. To examine this further, we used a spatial graph theoretical measure of the residual fast ripple rate-distance radius following a resection. This measure uses a combination of fRonS and fRonO > 350 Hz rates (events/min) and weights them by the spatial distance between the generator sites. Using this measure we found significantly lower residual fast ripple rate-distance in the 10 seizure free patients (ranksum, p<0.05, Figure 5E). Because our results show the SIZ and SOZ^29^ had been resected in most of the subjects overall, but low levels of fast ripple-generating tissue were found primarily in the seizure free subjects, it can be concluded that while the SIZ is epileptogenic, epileptogenic regions can be found outside the SIZ and the SOZ and generate solitary fRonO.

## Discussion

Bridging prior work demonstrating that HFOs preceding spikes have increased neuronal firing rates^38^, and propagating fRonO increase the probability of an after-going spike^26^, this study shows that the coincidence of fRonO preceding (<300 ms) spikes is statistically more common than chance, and that fRonO preceding spikes correspond with increase neuronal excitability, which could help generate an ensuing spike. We also found during the spike, neuronal firing was reduced, which could associate with an inhibitory restraint that has been demonstrated in preventing interictal spike propagation^39–42^. We extrapolated on these mechanistic findings in a larger cohort of patients who underwent resections and found that fRonO preceding spikes were generated in regions defined as the epileptogenic SOZ by the clinician that were subsequently resected, which supports fRonO preceding spikes demarcating a SIZ^9,37^. However, resecting the SIZ did not correlate with seizure free post-operative seizure outcome which suggests that epileptogenic regions are necessary but not sufficient for spike generation.

### Unit firing rate properties that underlie increased excitability changes during spike priming

A novel finding of our study was the strong correlation between increased spectral power of HFO and corresponding peak unit firing rates during the HFO. The HFO spectral power can reflect the recruitment of a larger pool of neurons for HFO generation in pathological regions^43^, although measurements of spectral power are confounded by differences in the distance from the generator to the electrode contact^44^. The fRonO preceding spikes exhibited higher associated firing rates than solitary fRonO while the corresponding spectral power of these fRonO events was much greater than solitary fRonO events. This suggests that a network effect involving macroscale neuronal recruitment is partly responsible. However, after controlling for increased fRonO power, the firing rates associated with pre-spike fRonO were still increased as compared to solitary fRonO. We also found that baseline firing rate was negatively correlated^7^ with the power of most forthcoming HFO types. A plausible explanation is that excitatory and inhibitory balance changes in neuronal ensemble activity, perhaps related to the UP and DOWN states, may also be predictive of a forthcoming HFO. We did not distinguish putative excitatory from inhibitory single units in this study but a prior investigation of the single unit correlates of epileptiform discharges found no difference in the firing properties of excitatory and inhibitory single units^7^. Together, these findings may help better explain the changes in network properties responsible for HFO and spike generation^17^ and spike priming by fRonO.

### Unit firing rate properties that may underlie inhibitory restraint after spike priming

We observed that the mean firing rate of the grand average of all fRonS and RonS event-unit AP trains more than doubled from baseline during the fRonS/ RonS. However, the overall firing rates were lower than that reported in similar investigations^7,17^. The most likely explanation is our use of a Gaussian kernel and a peak firing measure to estimate AP trains, which may also explain why we did not identify units that decreased in firing during HFO spike^7,17^. Importantly all the studies of the firing rate during interictal discharges measured from *in vivo* recordings in patients show that they are substantially less than that reported in human cortical slices^45^, and much less than in chemoconvulsant treated animals^46,47^.

In our study, when an fRonS/RonS followed a fRonO it exhibited decreased baseline AP firing before the event and decreased AP peak firing at the time of the event. We also observed, but did not quantify, that following the generation of all the HFO types, unit firing rate decreased for at least 500 msec. A recent study found that following fRonS generation a decreased in firing rates is seen even spatially far from the generator^48^. Thus, it is possible that decreased excitability during a fRonS/RonS is a result of the preceding fRonO or RonO alone. However, when RonS followed RonO decreased excitability was not observed in the baseline firing rate but was observed only during the RonS. This dissociation suggests that the decreased firing rate could not have resulted exclusively from RonO generation. Also, the decreased excitability during the fRonS following fRonO was more statistically significant than the decreased excitability seen during the baseline epoch preceding the fRonS. One interpretation is that decreased excitability during HFO spikes following RonO and fRonO results from an inhibitory restraint mechanism initiated during or just after priming. Inhibitory restraint of spike propagation has already been well established in animal models^39–42^, but its importance in spike initiation is less clear.

Although evidence for inhibitory restraints was seen for RonS followed by a RonO, we think it is more likely that fRonO prime spikes and less often RonO, because RonO preceded spikes less than chance, and exhibited no associated changes in excitability when they preceded spikes. Together, the results imply that HFO spikes that follow fRonO exhibit decreased excitability due to an inhibitory restraint mechanism functioning to prevent spike initiation.

### The role of fRonO propagation in spike priming

We previously found that fRonO that propagate in the SOZ are strongly coupled to the DOWN state and are more often followed by epileptiform discharges than non-propagating fRonO. In this case the fRonO precede the epileptiform spikes by <10 ms^26^. Here we could not relate fRonO propagation to unit firing activity because iEEG recordings were only available from the mesial most electrodes. However, we found that fRonO that precede epileptiform spikes by <10 ms may be distinct from fRonO that precede spikes by 10-300 ms (Fig S1,S2). Thus, based on the current results we can elaborate on our past results and propose a fRonO-spike complex, in which the fRonO precedes the spike by <10 ms, can propagate together, but an earlier event >10 ms may be required to prime this complex to occur first.

### Epileptogenic regions are necessary but not sufficient for spike priming

The clinical gold standard for planning resective epilepsy surgery is electroclinical correlation and removal of the SOZ^49^. In our clinical cohort, we found that fRonO preceding spikes were almost always generated in the SOZ. Which, according to our analysis of the unit activity corresponding to the paired events, indicates that the SIZ strongly overlapped with the SOZ (Figure 6). Moreover, only in the SOZ did fRonO precede spikes at rates greater than chance. Past work has shown that spikes following gamma oscillation (30-100 Hz) bursts are more exclusively generated in either the SOZ, or in resected regions in seizure free patients than spikes alone^50,51^. It is likely fRonO priming is not the sole mechanism for spike initiation, and gamma oscillations, that occur very frequently, also play a role and may generate different types of spikes.

In addition to fRonO preceding spikes occurring primarily within the SOZ, fRonO preceding spikes were also mainly generated at sites within the resection margins, in both the patients who were rendered seizure free by surgery, and the patients with failed surgery. This implies that for the patients with failed surgeries, other epileptogenic regions outside of the SOZ and SIZ could promote seizures after the surgery. Our data show that this other region, the residual epileptogenic zone (EZ)^13,49,52^, maybe identified using spatial estimates of residual FR generation (Figure 6)^29^ and such a region may generate solitary fRonO and seizures beyond its boundaries. Our results do not imply that the SIZ can be left intact and a seizure free outcome still achieved. To the contrary, our past investigation of intraoperative iEEG recorded from subdural electrodes in temporal lobe epilepsy patients found that unresected fRonS^22^, a strong biomarker of the SIZ, correlates with failed surgery. In addition, earlier studies had found that patients with well-localized spikes^53,54^, and spikes that post-operatively strongly decreased in occurrence, are more likely to be seizure free^53–55^. However, intraoperative electrocorticography which involves resecting regions using spike rates measured in subdural electrodes alone does not always result in seizure freedom, even when a combination of ripple and FR are used instead^56,57^.

A limitation of our study is that source localization, employing the temporal ordering of discharges, was not used to identify the SIZ because it required distinct methods for isolating bluntly contoured discharges. Two studies implementing this method similarly localized the SIZ to epileptogenic regions^9,37^, although this was not the case in another study utilizing microelectrode arrays instead of macroelectrodes^8^. It will be necessary in the future to combine fRonO and spike coincidence analysis with iEEG spike source localization analysis to determine if the location of the SIZ is agreed upon.

## Conclusion

We show that the generation of a fRonO, reflecting increased neuronal excitability, primes the generation of an after-going spike within an epileptogenic region. Additionally, during the initiation of this subsequent spike, excitability is reduced relative to solitary spikes. This likely reflects an inhibitory restraint that is well established for preventing the spatial propagation of spikes. In a clinical context, our results show that epileptogenic regions are necessary but not sufficient for spike generation. This is important in current practice because strategies to localize and resect just the SIZ or SOZ alone may not always succeed because of the possibility of a residual EZ. In future practice it may be possible to prevent both spikes and seizures by pharmacologically or electrically inhibiting the occurrence of fRonO.

## Data Availability

The data and code to reproduce the results and figures are available at https://zenodo.org/record/7685943#.Y_5UHybML9Z and https://github.com/shenweiss/FRexcitabilityprespike. The raw data is available upon reasonable request.

https://zenodo.org/record/7685943#.Y_5UHybML9Z

## Acknowledgements

The authors thank Dr. Chenyuan Wu and Dr. Ashwini Sharan for their neurosurgical contributions, Mr. Kirk Shattuck, Dr. Iren Orosz, and Mr. Dale Wyeth and Ted Wyeth for their technical contribution in collecting EEG recordings, and Dr. Iren Orosz and Dr. Richard Gorniak for assistance with the neuroimaging data. We would also like to thank Christina Louise George Trust and the Resnick Family Foundation.

## Contributions

Shennan A. Weiss: Conceptualization, Methodology, Software, Investigation, Resources, Writing – original draft, Writing – review & editing, Funding acquisition. Itzhak Fried: Resources. Jerome Engel: Writing – review & editing, Funding acquisition. Michael R. Sperling: Writing – review & editing, Funding acquisition. Yuval Nir: Conceptualization, Methodology, Software, Investigation, Resources, Writing – review & editing. Richard J. Staba: Conceptualization, Methodology, Writing – review & editing, Funding acquisition.

