## supplemental materials for "Fast ripples reflect increased excitability that primes epileptiform spikes"

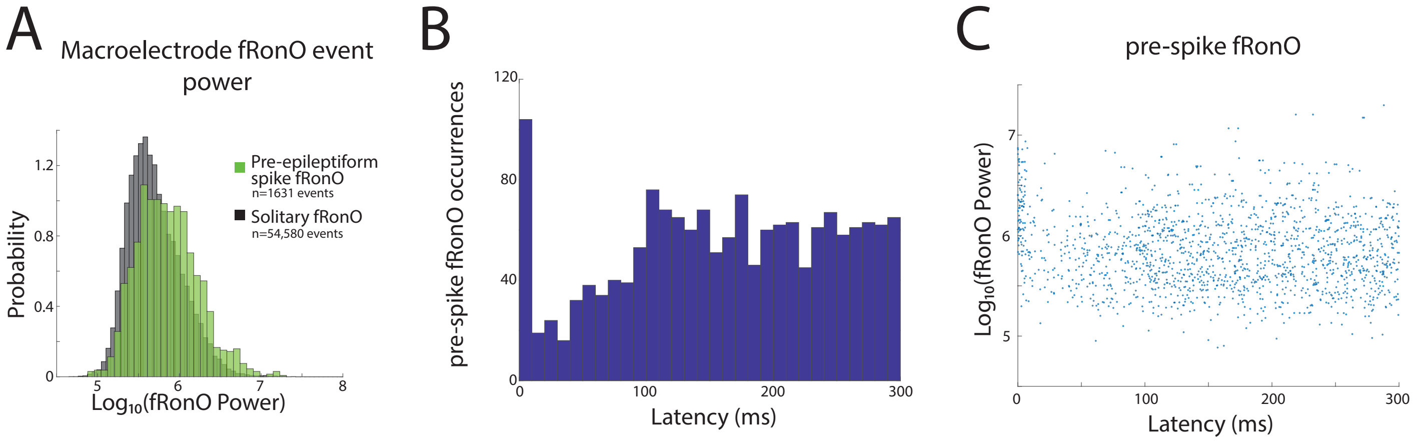


Figure S1: An examination pre-epileptiform spike fast ripple on oscillation (fRonO) power with increased temporal resolution. (A) Normalized histogram of fRonO event spectral power. fRonO events preceding spikes had a larger power (t-test, p<1e-10, Cohen's d=.475) than solitary fRonO. (B) Histogram of the latency in milliseconds (ms) between pre-spike fRonO and the after-going spike. Note that a relatively small, but distinct, subpopulation of events occurred within <10 msec of the spike. (C) Plot of the pre-spike fRonO power as a function of latency. Note that relatively larger powers (see A) were seen across a broad range of latencies. However, fRonO events occurring <10 ms before the after-going spike had the largest power distribution

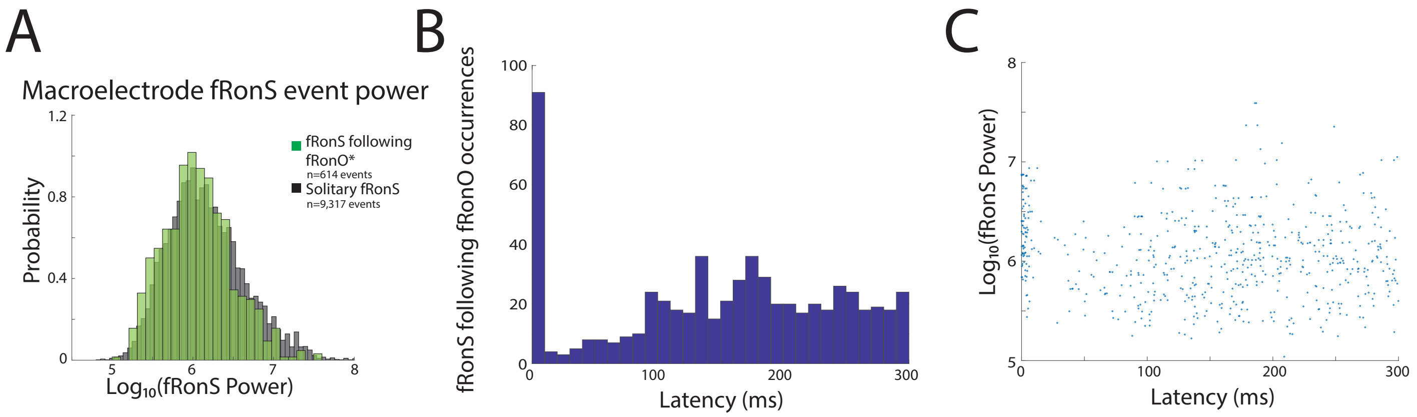


Figure S2: An examination of fast ripple on spike (fRonS) following fast ripple on oscillation (fRonO) power with increased temporal resolution. (A) Normalized histogram of fRonS event power in macroelectrode recordings. fRonS power was reduced when it followed a fRonO (t-test, p<1e-5, Cohen's d=0.21) (B) Histogram of the latency in milliseconds (ms) between the fRonO and the after-going fRonS. Note that a distinct subpopulation of events occurred within <10 msec of the spike. (C) Plot of the fRonS following fRonO power as a function of latency. Note that relatively decreased powers (see A) were seen across a broad range of latencies. However, fRonS events following a fRonO by <10 ms had the largest power distribution suggesting a distinct mechanism.

Table S1: Patient characteristics in the resection Table S4: Results of generalized linear mixed effects model for the outcome of fast ripple (FR) power (arbitrary units A.U.) with random effect term of electrode contact (i.e., node) pair (i.e., edge) number. The fixed effects of the model were 1) whether the fast ripple was recorded from the out-node or in-node (abbr., inout); 2) whether the fast ripple was considered propagating from the sign test either from the out-node (abbr., prop); 3) the Euclidian distance of propagation between the in- and out nodes; 4) whether the out- or in-node was located in the seizure-onset zone (abbr., soz); d.f.=39,001. Brackets indicate

| ID | Risk Factor | MRI | PET (hypo-metabolic) | iEEG clinical consensus SOZs | Surgery | Path. | Outcome |
|---|---|---|---|---|---|---|---|
| 1. | minor TBI | Normal | L tempo-ral | L MT | modified L ATL hippocampus sparing | Gliosis | Engel IA@24 months |
| 2. | hyperten-sive enceph-alopathy | post L ATL | N/A | L middle tem-poral gyrus | modified L temporal lo-bectomy (posterior tem-poral included) | Gliosis | Engel 1A @48 months |
| 3. | Minor TBI | Normal | Normal | Right insula, cuneus, inferi-or and middle frontal gyrus | R. Frontal lobe | Gliosis | Engel IA@24 months |
| 4. | None | Normal | R temporal | R Inferior tem-poral gyrus | modified R temporal lo-bectomy (posterior tem-poral included) | Gliosis | Engel IA@24 months |
| 5. | None | L MTL whitematter hyper-inten-sity | Normal | R SMA | R frontal lobe resection | cortical dyspla-sia | Engel IA@24 months |
| 6. | None | Normal | Normal | L cingulate gyrus, medial frontal gyrus, middle frontal gyrus, superior frontal gyrus | L frontal lobec-tomy | cortical dyspla-sia | Engel IA@40 months |
| 7. | meningitis | Encepha-lo-malacia | L temporal | L MT, uncus, superior tem-poral gyrus, frontal lesion | L temporal and frontal lobe resection | gliosis | Engel IB@42 months |
| 8. | none | 1 cm pineal cyst | R lateral temporal | L MT | Modified L temporal lo-bectomy (pos-terior temporal included) | Gliosis | Engel IIB@24 months |

| | | | | | | | |
|---|---|---|---|---|---|---|---|
| 9. | None | T2 hyper-intensity in R temporal pole > L frontal pole. Inferior portion of R temporal pole with blurred gray-white matter border | R temporal | R MT | R anterior ATL | Cortical dysplasia IIb | Engel IA@60 months |
| 10. | None | Normal | R temporal | Bilateral MT, middle temporal gyrus R>L | modified R ATL (preserved middle and superior temporal gyrus) | gliosis | Engel IVC@48 months |
| 11. | TBI, family history | left superior temporal gyrus encephalomalicia | L parieto-occipital | L temporal neocortical, L frontal | modified LATL hippocampus sparing | gliosis | Engel IV@6 months RNS placed and revised |
| 12. | None | Normal | L temporal | R fusiform gyrus, superior temporal gyrus, uncus | R ATL | MTS | Engel IB@35 months |
| 13. | TBI w/ LOC | L MTS, extra-temporal T2 | L temporal and frontal | L MT, fusiform gyrus, uncus | L MT visualase | N/A | Engel IIIA@18 months |
| 14. | None | periventricular nodular heterotopia, right frontal T2 | R temporal | R MT | ATL | gliosis | Engel IB@31 months |
| 15. | TBI w/ LOC | Encephalomalacia | R temporal | R insula, bilateral middle temporal gyrus, superior temporal gyrus | modified R ATL (posterior temporal and temporal-parietal-occipital junction included. | gliosis | Engel IVB@33 months |

| | | | | | | | |
|---|---|---|---|---|---|---|---|
| 16. | None | Prior R. ATL | N/A | Right orbitofrontal cortex. | R. Frontal lobe | hippocampal sclerosis, cortical dysplasia | Engel IVB@24 months |
| 17. | encephalitis | Encephalomalacia | Normal | L inferior frontal gyrus, insula, MT | L temporal lobe and insula resection | Gliosis | SUDEP @6 weeks |
| 18. | Significant head injury with LOC | Left temporal T2 hyperintensity with mild enhancement | N/A | Bilateral MT, right lateral temporal | L temporal lobectomy, anterior thalamic DBS | Gliosis | Engel IVB@24 months |
| 19. | None | R parietal lobe resection | R parietal and R occipital | R insula, precuneus, middle occipital gyrus, superior parietal lobule, superior occipital gyrus, superior temporal gyrus, middle temporal gyrus | R parietal | gliosis | Engel IIIA@18 months |
| 20. | None | L posterior fossa arachnoid cyst, R ATL | R temporal | L MT, R cingulate, post. cingulate, mesial frontal, precuneus | R anterior cingulate thermal ablation | gliosis | Engel IVB@36 months |
| 21. | None | Prior R parietal resection | R parietal and occipital hypometabolism | R parietal lobe | R. Parietal lobe resection | Gliosis | Engel IVB@36 months |
| 22 | TBI | left MTS | normal | L MT | L ATL | gliosis hippocampal sclerosis | Engel IIIA@63 months |
| 23 | febrile seizures | prior hippocampal sparing temporal lobectomy | N/A | R anterior cingulate, MT, uncus | R ATL | gliosis | Engel IVB@40 months |

Table S2: Results of generalized linear mixed effects model for the outcome of the maximum unit firing rate during the fast ripple on oscillation (fRonO) detected in the macroelectrode, (action potentials/sec) after Gaussian smoothing, minus the mean baseline unit firing rate measured 750 msec before the fRonO for each peri-fRonO trial (d.f.=224,266). The random effect terms were patient id, macroelectrode contact ID of the Behnke-Fried electrode, and the unit id to determine that the effects were observed across all units. The fixed effects of the model were 1) the log10(power) of the trial's fRonO measured in arbitrary units, and 2) whether, in the trial, the fRonO preceded a sharply contoured epileptiform spike, with or without a high-frequency oscilation, within 300 msec. ":" refers to interaction term. Abbreviations CI: confidence interval.

| fRonO unit firing rate - baseline firing rate<br>Fixed effect name | Estimate [95% CI] | tStat | p-value |
|---|---|---|---|
| (intercept) | -8.528 [-9.012 -8.043] | -34.472 | <1e-100 |
| 1. power | 1.044 [0.997 1.089] | 46.026 | <1e-100 |
| 2. preSpike | 1.740 [0.764 2.716] | 3.493 | <1e-3 |
| 3. preSpike:power | -0.233 [-0.392 -0.734] | -2.863 | <1e-2 |

Table S3: Results of generalized linear mixed effects model for the outcome of the mean baseline unit firing rate measured 750 msec before the fast ripple on oscillation (fRonO) for each peri-fRonO trial (d.f.=224,266). The random effect terms were patient id, macroelectrode contact ID of the Behnke-Fried electrode, and the unit id to determine that the effects were observed across all units. The fixed effects of the model were 1) the log10(power) of the trial's fRonO measured in arbitrary units, and 2) whether, in the trial, the fRonO preceded a sharply contoured epileptiform spike, with or without a high-frequency oscilation, within 300 msec. ":" refers to interaction term. Abbreviations CI: confidence interval.

| pre-fRonO<br>baseline unit firing rate | Estimate [95% CI] | tStat | p-value |
|---|---|---|---|
| (intercept) | -1.326 [-1.680 -0.972] | -7.353 | <1e-12 |
| 1. power | -0.030 [-0.040 -0.019] | -5.426 | <1e-7 |
| 2. preSpike | 0.145 [-0.086 0.381] | 1.240 | n.s. |
| 3. preSpike:power | -0.023 [-0.063 0.017] | -1.141 | n.s. |

Table S4: Results of generalized linear mixed effects model for the outcome of the maximum unit firing rate during the ripple on oscillation (RonO) detected in the macroelectrode, (action potentials/sec) after Gaussian smoothing, minus the mean baseline unit firing rate measured 750 msec before the RonO for each peri-RonO trial (d.f.=2,603,844). The random effect terms were patient id, macroelectrode contact ID of the Behnke-Fried electrode, and the unit id to determine that the effects were observed across all units. The fixed effects of the model were 1) the log10(power) of the trial's RonO measured in arbitrary units, and 2) whether, in the trial, the RonO preceded a sharply contoured epileptiform spike, with or without a high-frequency oscilation, within 300 msec. ":" refers to interaction term. Abbreviations CI: confidence interval.

| RonO unit firing rate - baseline firing rate<br>Fixed effect name | Estimate [95% CI] | tStat | p-value |
|---|---|---|---|
| (intercept) | -7.909 [-8.367 -7.450] | -33.823 | <1e-100 |
| 1. power | 0.800 [0.788 0.811] | 131.56 | <1e-100 |
| 2. preSpike | -0.514 [-0.822 -0.206] | -3.270 | <5e-3 |
| 3. preSpike:power | 0.095 [0.0501 0.139] | 4.170 | <1e-4 |

Table S5: Results of generalized linear mixed effects model for the outcome of the mean baseline unit firing rate measured 750 msec before the ripple on oscillation (RonO) for each peri-RonO trial (d.f.=2,603,845). The random effect terms were patient id, macroelectrode contact ID of the Behnke-Fried electrode, and the unit id to determine that the effects were observed across all units. The fixed effects of the model were 1) the log10(power) of the trial's RonO measured in arbitrary units, and 2) whether, in the trial, the RonO preceded a sharply contoured epileptiform spike, with or without a high-frequency oscilation, within 300 msec. ":" refers to interaction term. Abbreviations CI: confidence interval.

| pre-RonO<br>baseline unit firing rate | Estimate [95% CI] | tStat | p-value |
|---|---|---|---|
| (intercept) | -1.858 [-2.123 -1.592] | -13.729 | <1e-42 |
| 1. power | -0.020 [-0.024 -0.017] | -10.495 | <1e-25 |
| 2. preSpike | -0.096 [-0.177 -0.014] | -2.296 | <0.05 |
| 3. preSpike:power | 0.016 [0.004 0.028] | 2.568 | <0.05 |

Table S6: Results of generalized linear mixed effects model for the outcome of the maximum unit firing rate during the fast ripple on spike (fRonS) detected in the macroelectrode, (action potentials/sec) after Gaussian smoothing, minus the mean baseline unit firing rate measured 750 msec before the fRonS for each peri-fRonS trial (d.f.=48,164). The random effect terms were patient id, macroelectrode contact ID of the Behnke-Fried electrode, and the unit id to determine that the effects were observed across all units. The fixed effects of the model were 1) the log10(power) of the trial's fRonS measured in arbitrary units, and 2) whether, in the trial, the fRonS followed a fast ripple on oscillation (fRonO) within 300 msec. ":" refers to interaction term. Abbreviations CI: confidence interval.

| fRonS unit firing rate - baseline firing rate<br>Fixed effect name | Estimate [95% CI] | tStat | p-value |
|---|---|---|---|
| (intercept) | -1.887 [-2.42 -1.34] | -6.846 | <1e-11 |
| 1. power | 0.125 [0.105 0.144] | 12.464 | <1e-34 |
| 2. followingFRonO | -2.830 [-3.662 -2.0] | -6.657 | <1e-10 |
| 3. followingFRonO:power | 0.430 [0.301 0.559] | 6.549 | <1e-10 |

Table S7: Results of generalized linear mixed effects model for the outcome of the maximum unit firing rate during the fast ripple on spike (fRonS) detected in the macroelectrode, (action potentials/sec) after Gaussian smoothing, minus the mean baseline unit firing rate measured 750 msec before the fRonS for each peri-fRonS trial (d.f.=48,164). The random effect terms were patient id, macroelectrode contact ID of the Behnke-Fried electrode, and the unit id to determine that the effects were observed across all units. The fixed effects of the model were 1) the log10(power) of the trial's fRonS measured in arbitrary units, and 2) whether, in the trial, the fRonS followed a ripple on oscillation (RonO) within 300 msec. ":" refers to interaction term. Abbreviations CI: confidence interval.

| fRonS unit firing rate - baseline firing rate Fixed effect name | Estimate [95% CI] | tStat | p-value |
|---|---|---|---|
| (intercept) | -2.019 [-2.565 -1.473] | -7.250 | <1e-12 |
| 1. power | 0.141 [0.120 0.162] | 13.14 | <1e-38 |
| 2. followingRonO | 0.307 [0.048 0.565] | 2.34 | <0.05 |
| 3. followingRonO:power | -0.036 [-0.077 0.005] | 1.73 | n.s. |

Table S8: Results of generalized linear mixed effects model for the outcome of the mean baseline unit firing rate measured 750 msec before the fast ripple on spike (fRonS) for each peri-fRonS trial (d.f.=48,164). The random effect terms were patient id, macroelectrode contact ID of the Behnke-Fried electrode, and the unit id to determine that the effects were observed across all units. The fixed effects of the model were 1) the log10(power) of the trial's fRonS measured in arbitrary units, and 2) whether, in the trial, the fRonS followed a fast ripple on oscillation (fRonO) within 300 msec. Abbreviations CI: confidence interval.

| pre-fRonS<br>baseline unit firing rate | Estimate [95% CI] | tStat | p-value |
|---|---|---|---|
| (intercept) | -1.408 [1.902 -0.914] | -5.589 | <1e-7 |
| 1. power | -0.0123 [-0.024 -0.001] | -2.105 | <0.05 |
| 2. followingFRonO | -0.742 [-0.896 -0.049] | -2.186 | <0.05 |
| 3. followingFRonO:power | 0.0720 [0.004 0.14] | 2.062 | <0.05 |

Table S9: Results of generalized linear mixed effects model for the outcome of the mean baseline unit firing rate measured 750 msec before the fast ripple on spike (fRonS) for each peri-fRonS trial (d.f.=48,164). The random effect terms were patient id, macroelectrode contact ID of the Behnke-Fried electrode, and the unit id to determine that the effects were observed across all units. The fixed effects of the model were 1) the log10(power) of the trial's fRonS measured in arbitrary units, and 2) whether, in the trial, the fRonS followed a ripple on oscillation (RonO) within 300 msec. Abbreviations CI: confidence interval.

| pre-fRonS<br>baseline unit firing rate | Estimate [95% CI] | tStat | p-value |
|---|---|---|---|
| (intercept) | -1.392 [-1.887 -0.900] | -5.519 | <1e-7 |
| 1. power | -0.017 [-0.030 -0.003] | -2.452 | <0.05 |
| 2. followingRonO | -0.103 [-0.247 0.040] | -1.408 | n.s. |
| 3. followingRonO:power | 0.022 [-0.0004 0.044] | 1.918 | n.s. |

Table S10: Results of generalized linear mixed effects model for the outcome of the maximum unit firing rate during the ripple on spike (RonS) detected in the macroelectrode, (action potentials/sec) after Gaussian smoothing, minus the mean baseline unit firing rate measured 750 msec before the RonS for each peri-RonS trial (d.f.=391,663). The random effect terms were patient id, macroelectrode contact ID of the Behnke-Fried electrode, and the unit id to determine that the effects were observed across all units. The fixed effects of the model were 1) the log10(power) of the trial's RonS measured in arbitrary units, and 2) whether, in the trial, the RonS followed a fast ripple on oscillation (fRonO) within 300 msec. ":" refers to interaction term. Abbreviations CI: confidence interval.

| RonS unit firing rate - baseline firing rate Fixed effect name | Estimate [95% CI] | tStat | p-value |
|---|---|---|---|
| (intercept) | -3.272 [-3.693 -2.852] | -15.253 | <1e-51 |
| 1. power | 0.260 [0.250 0.270] | 50.411 | <1e-100 |
| 2. followingFRonO | -2.684 [-3.225 -2.144] | -9.738 | <1e-21 |
| 3. followingFRonO:power | 0.372 [0.298 0.445] | 9.862 | <1e-22 |

Table S11: Results of generalized linear mixed effects model for the outcome of the maximum unit firing rate during the ripple on spike (RonS) detected in the macroelectrode, (action potentials/sec) after Gaussian smoothing, minus the mean baseline unit firing rate measured 750 msec before the RonS for each peri-RonS trial (d.f.=391,663). The random effect terms were patient id, macroelectrode contact ID of the Behnke-Fried electrode, and the unit id to determine that the effects were observed across all units. The fixed effects of the model were 1) the log10(power) of the trial's RonS measured in arbitrary units, and 2) whether, in the trial, the RonS followed a ripple on oscillation (RonO) within 300 msec. ":" refers to interaction term. Abbreviations CI: confidence interval.

| RonS unit firing rate - baseline firing rate<br>Fixed effect name | Estimate [95% CI] | tStat | p-value |
|---|---|---|---|
| (intercept) | -3.183 [-3.603 -2.763] | -14.861 | <1e-49 |
| 1. power | 0.245 [0.236 0.259] | 42.905 | <1e-100 |
| 2. followingRonO | -0.535 [-0.691 -0.380] | -6.733 | <1e-10 |
| 3. followingRonO:power | 0.076 [0.055 0.098] | 6.923 | <1e-11 |

Table S12: Results of generalized linear mixed effects model for the outcome of the mean baseline unit firing rate measured 750 msec before the fast ripple on spike (RonS) for each peri-RonS trial (d.f.=391,663). The random effect terms were patient id, macroelectrode contact ID of the Behnke-Fried electrode, and the unit id to determine that the effects were observed across all units. The fixed effects of the model were 1) the log10(power) of the trial's RonS measured in arbitrary units, and 2) whether, in the trial, the RonS followed a fast ripple on oscillation (fRonO) within 300 msec. Abbreviations CI: confidence interval.

| pre-RonS<br>baseline unit firing rate | Estimate [95% CI] | tStat | p-value |
|---|---|---|---|
| (intercept) | -1.510 [-1.935 -1.084] | -6.956 | <1e-11 |
| 1. power | -0.026 [-0.031 -0.022] | -11.119 | <1e-27 |
| 2. followingFRonO | -0.671 [-0.846 -0.495] | -7.499 | <1e-13 |
| 3. followingFRonO:power | 0.099 [0.743 0.124] | 7.825 | <1e-14 |

Table S13: Results of generalized linear mixed effects model for the outcome of the mean baseline unit firing rate measured 750 msec before the fast ripple on spike (RonS) for each peri-RonS trial (d.f.=391,663). The random effect terms were patient id, macroelectrode contact ID of the Behnke-Fried electrode, and the unit id to determine that the effects were observed across all units. The fixed effects of the model were 1) the log10(power) of the trial's RonS measured in arbitrary units, and 2) whether, in the trial, the RonS followed a ripple on oscillation (RonO) within 300 msec. Abbreviations CI: confidence interval.

| pre-RonS baseline unit firing rate | Estimate [95% CI] | tStat | p-value |
|---|---|---|---|
| (intercept) | -1.544 [-1.968 -1.120] | -7.135 | <1e-12 |
| 1. power | -0.023 [-0.029 -0.018] | -8.499 | <1e-16 |
| 2. followingRonO | -0.014 [-0.080 0.053] | -0.402 | n.s. |
| 3. followingRonO:power | 0.010 [0 0.020] | 2 | n.s. |
